# Challenges of stakeholders’ engagement for developing pragmatic, primary health care interventions for chronic respiratory diseases in low resource settings in India

**DOI:** 10.1101/2022.04.15.22272333

**Authors:** Rita Isaac, R Harsh, Biswajit Paul, David Weller, Paul Jebaraj, Bochu Vikas, BV Balaji, Surenthiran Natarajan, Tracy Jackson, Genevie Fernandes, RESPIRE collaborators

## Abstract

Chronic respiratory diseases (CRDs) in low resource settings in India are often poorly diagnosed, leading to missed opportunities for early initiation of treatment and poor patient pathways. There is also a poor understanding in rural communities of the causes of CRDs; many symptoms are incorrectly attributed to ‘asthma’ and treatment is inconsistent and often based on inaccurate diagnoses. There is a high prevalence of CRDs in rural regions of India. It is vital that interventions are developed to improve an understanding about CRD in low resource settings in India in order to reduce exposure to common risk factors and improve access to evidence based care. We piloted a frontline health care worker delivered ‘educate, screen and treat’ intervention programme. We explored the role of stakeholders’ engagement towards the development of feasible community based interventions. The hypothesis was that without meaningful engagement with the key stakeholders, long term sustainability of the programme would be limited and potentially viewed as primarily for the organisation’s self-interest. A mixed method study combined a quantitative online survey of the sensitised health care providers and a qualitative assessment of the other stakeholders’ engagement activities. The methods of qualitative data collection included focus group discussions, feedback and individual interviews and data analysis used a thematic framework. We identified key stakeholders and investigated their knowledge, perceptions, beliefs, practices, educational needs and suggestions for improved care for CRD. Three main themes were 1) Community trust building 2) Mismatch between community awareness about CRD and access to evidence based care resources 3) Finding effective communication methods for low health literate and older age group population with CRD. First theme was built on two sub-themes: sensitised influential people in the community (community advisory committee); empowered, trained health workers in the community for patient screening and navigation. Second theme informed by three sub-themes: availability of sensitized health care providers and empowered health system; recognizing access to evidence based care for CRD in the region/district; recognizing the community’s change in behavior related to management of CRD with education. Third theme was informed by 2 sub-themes: community’s ability to understand the messages through different educational media and tools; challenges in engaging the low health literacy population. The findings of this study add to the literature on the numerous challenges faced by patients with CRD in low resource settings, indicating the need for identifying and educating key stakeholders, continuous support of patients, personalised education, and capacity building of health care providers. Given the significant challenges that the patients face, a feasible primary health care model needs to be developed incorporating strategies to deal with these challenges.

## Background

Chronic Respiratory Diseases (CRDs), including asthma, chronic obstructive pulmonary disease (COPD) and bronchiectasis, are common public health problems with high prevalence and mortality rates globally and more so in developing countries. ^1,2^ COPD is the 3rd leading cause of death in the world; around 90% of all COPD deaths and 88% of all disability-adjusted life years (DALYs) due to COPD occur in developing countries. ^3^ Asthma is the 16^th^ most important disorder in the world in terms of the extent and duration of disability.^4^ It is estimated that asthma alone accounts for the loss of approximately 15 million DALYs and 180,000 deaths annually representing approximately 1% of the total global disease burden, and its prevalence has been increasing by approximately 50% every decade.^5^ This study health center reported about 15 percent of the out-patient attendance in 2016-17 was for CRDs.^6^

In low resource settings, CRDs are often underdiagnosed, leading to missed opportunities for early initiation of treatment and poor patient pathways.^7,8^ Further it has been observed that the compliance to treatment for recommended longer duration is poor due to the limited knowledge of the causes of respiratory illness, symptoms and treatment options among people. Interestingly, any symptoms of CRD are often incorrectly attributed to ‘asthma’ by health care providers, resulting in inappropriate treatment.^9,10^ It is vital that interventions are developed to improve an understanding about CRD in low resource settings in India to change risk behaviors, reduce exposure to common risk factors (particularly smoking and exposure to indoor cooking smoke) and improve access to evidence based treatment. We, in partnership with the NIHR Global Health Research Unit on Respiratory Health (RESPIRE) at University of Edinburgh, piloted a frontline health care worker delivered ‘*educate, screen and treat’* intervention program for CRD in a resource limited rural setting in India. The study was done during June 2018 to March 2021. Towards the development of culturally appropriate, feasible, community based interventions to address issues related to management of CRD in the rural areas, we explored the role of stakeholder engagement. We undertook significant stakeholder engagement activities to identify the best tools for effective communication within the available resources, foster trust and confidence of communities, and support for our new intervention on CRD care. We hypothesized that without meaningful engagement with the key stakeholders, long term sustainability of the program would be limited and potentially viewed as primarily for the organization’s self-interest. The World Health Organization (WHO) defines stakeholder engagement as the process of developing relationships that enable stakeholders to work together to address health-related issues and promote well-being to achieve positive health outcomes.^11^ In this paper, we present the process of identifying stakeholders, methods of engagement, stakeholders’ perceptions and experiences of the factors influencing implementation and patient participation in the program.

## Materials and methods

### Study setting

The study was undertaken in a rural population in the State of Tamil Nadu, India. The residents of the study villages were mostly engaged in agriculture related jobs; other occupations include small businesses, weaving, poultry farming and salaried service jobs. About 97% of the population identified as Hindus; literacy level among men was approximately 74% and among women was 57%; sex ratio was 984; ∼30% of the households use biomass fuel as an alternative source of energy for cooking. A recent study conducted by the investigators showed a CRD prevalence rate of 14 percent in this population.^12^

This study obtained institutional review board approval from Christian Medical College, Vellore, India (IRB Min. No. 11381 dated 27/06/2018). A written consent was taken from all the participants after informing them about the purpose and methods of the study.

### Selection of stakeholders and methods of engagement

The selected stakeholders and their roles in the feasibility of intervention study are outlined in figure 1.

**Fig. 1.** Stakeholders’ role to provide evidence based care for Chronic Respiratory Diseases. The first step of stakeholders’ engagement was to identify those stakeholders who can promote the proposed primary health care based interventions and make suggestions for improvement. We had identified primary care physicians in the district for whom a workshop was organized prior to project inception, followed by a continuing medical education (CME) workshop; the primary care nurses who attended a nursing conference were engaged through a talk and exhibition; and the health care workers who were expected to do the patient screening and navigate them for care were given training. The next step was to engage those who are impacted by CRD (patients, family members and community) to raise awareness, support behavior change, maximize screening uptake and improve compliance to treatment and follow-up. The community engagement used theory of planned behavior informed approach and the methods comprised of face-to-face, individual and group-based behaviour change interventions using leaflets, video-shows, live puppet shows, ‘health mela’(health festival) and school health programme. In addition, we formed a community advisory committee comprising of CRD patients, village leaders and key influencers in the community to ensure community participation in the development of the programme.

### Evaluation research design

The focus of the overall research was to develop feasible, effective interventions to enhance the ability of patients, providers and policymakers to make evidence-based decisions on CRD management in low resource settings. A mixed method study was conducted using quantitative online survey for a sample of sensitised healthcare providers, 3 months after the CME and qualitative assessment of the other stakeholders’ engagement activities. The qualitative study was based on grounded theory that involved the collection and analysis of actual data and the methods included focus group discussions, feedback, and individual interviews.^13^ We stopped the qualitative research after a theoretical saturation was reached at a point where we had sampled and analyzed the data and uncovered all data.^14^ The study explored the knowledge, change in attitude and practices regarding CRD management among the health care providers.

The community education was based on pragmatic, theory-of planned behavior informed approach. Community members who participated in the puppet shows, video shows, and received leaflets were assessed to gather the depth of understanding of the health messages. Individual, semi-structured interviews were conducted with the purposively sampled participants of the educational activities. Structured topic guides were developed for interviews. Interviews conducted in vernacular language (Tamil) were recorded, transcribed verbatim and translated into English by the transcribers and translators (PJ, BBV). Analysis employed the thematic framework method.

### Data analysis

The quantitative data from survey of health care providers was analysed using frequencies and percentages. Analysis of the qualitative data was conducted by a team of experts comprised of implementation research expert, community health physicians and a public health specialist who have had training in qualitative data analysis. Transcripts were manually analyzed using qualitative thematic framework analysis. Data analysis proceeded in two steps. (a) In the initial step, coding of the transcripts was carried out that described the CRD related knowledge acquired, perceived preventive measures, change of attitude, change in practices and behavior made, access to care and suggestions to ways of reaching community. This first step was conducted separately by H and PJ guided by RI. In the second step, preliminary codes were compared across the types of participants by RI, H and similar codes were grouped and put under the same categories and sub-categories which were later discussed and revised together with RI, H, and PJ during a series of meetings. By sorting the codes into categories, we were able to detect consistent and overarching themes emerging from the data and the sub-categories were the supporting/subthemes.

In order to enhance credibility and trustworthiness, H, BBV and PJ examined the transcripts from the interviews with the community members and the other stakeholders and together the data collection team reached a consensus on the final analysis. The process of thematic analysis is displayed in table. 1.

**Table1.** Themes and sub-themes.

| Theme | Subthemes |
| --- | --- |
| 1. Community trust building | a) Sensitised key people/influencers in the community (community advisory committee)<br>b) Empowered, trained Health workers in the community for patient screening and navigation |
| 2. Mismatch between community awareness about CRD and access to evidence based care resources | a) Availability of sensitized health care providers and empowered health system ; Recognizing access to evidence based care for CRD in the region/district<br>b) Recognizing the community's change in behavior related to management of CRD with education |
| 3. Finding effective communication methods for low health literate and older age group population with CRD. | a) Community's ability to understand the messages through different educational media and tools; Challenges in engaging the low health literacy population |

## Results

### 1. Community trust building

#### 1a. Sensitised key people/influencers in the community (community advisory committee)

In order to develop community trust, key people in the community were invited to form a community advisory group. They were expected to promote the proposed community interventions and obtain feedback from the wider community on the new CRD care initiative. Those who volunteered to participate were former ‘panchayat’(village council) presidents, coordinator and members of self-help women groups (SHG), village chiefs, an employee in the public works department, cooperative milk producers’ federation worker, a college student, farmers’ club members, people with CRD. They shared their perceived burden of CRD in the community, perceptions of cause and treatment of CRD and barriers to care.

Their comments and suggestions were:

One of the respondents said: *There are a lot of patients with breathlessness disease in our village, After they travel by bus to ‘RUHSA’ hospital and wait for many hours to collect medicine, their symptoms are worsened. If you arrange RUHSA Health aide to organize the clinic in the village health center to issue medications(a community pharmacy), everyone will come and get benefited; we see that you’re doing a very good project*

*We need to discuss about the increased CRD in people at the ‘Gram Sabha’ (a meeting of all adults/electorate who live in the area covered by a ‘Panchayat’) that is held monthly once at the village level. Village administrative officer and other government officials will be coming to the meeting and it’s the right place to discuss about having a dedicated place to burn the waste to reduce the smoke pollution; If we want to inform about the new CRD care initiative to the families, we need to educate the SHG women (self-help women group) and if we want to reach the community, we need to educate the ‘Gram Sabha’*.

*Some of the patients are feeling shy to tell they are having breathlessness disease and so they are not taking medicine; Some of them are near my home and they don’t know how to use the inhaler; a few of them are not able to take the inhaler as they don’t have the strength to take it; they are all working and not able to take care of their diseases; some don’t have money to buy the medicine*

They listed the causes of CRD as dust, smoking, cold foods, overweight, tobacco chewing, using firewood for cooking, burning solid waste and old age

#### 1b. Empowered, trained Health workers in the community for patient screening and navigation

The trained health workers were able to organize 77 educational video shows for the 100 study participants and their family members during the 1 year of the intervention; conduct once a month demonstration sessions on correct use of inhalers and respiratory exercises at the community centers close to the participants’ home and educate one-on-one using pamphlets (supplementary document). The four health workers over 6 month period screened 289 potential participants and out of them, 219 were assessed by the physician and 208 were confirmed to have CRD.

### 2. Mismatch between community awareness about CRD and access to evidence based care

#### 2a. Availability of sensitized providers and empowered health system and recognizing access to evidence based care for CRD in the region

A network of primary health care physicians who practice lung medicine in the district were brought together in a CME workshop on CRD management. Of the 80 participants who attended the CME, 21 responded to an online survey conducted 3 months after the workshop to assess the challenges they faced in providing evidence based care for CRD and what changes were made in their routine practice. The table.1 lists the challenges of providing evidence based care for CRD.

Though about 70% of the providers were comfortable using mMRC and GINA guidelines, about 38% of them did not have access to spirometer or peak flow meters to measure the lung function; 42.9% of them have changed the practice of using oral medications to recommended inhaler therapy. About 38% of the providers find it difficult to follow up the patients with clinical reviews.

#### 2b. Recognizing the community’s change in behavior related to CRD management with education

### 3. Finding effective communication methods for low health literate and older age group population with CRD

#### 3.a Community’s ability to understand the messages through different educational media and tools and challenges in engaging the low health literacy population

The people’s ability to understand the educational messages was as follows:

‘ *…. I don’t know to read. So I have got the handout and kept it with me. Health worker read it out for me, but I didn’t understand anything. From puppet show, I gathered those inhalers are important for my life. I need to do exercise such as walking and need to visit the doctor if I have severe problem in my breathing; I have seen the videos. I learned that we should not be near smoke, shouldn’t cook in ‘Chula’(earthen stove using firewood). Hmmm… we need to stop smoking ‘beedi’ and cigarette, inhalers are best for breathing difficulty than oral medicine’*. (Male, in his 70’s).

*From handout, I realized that I need to change fuel for cooking from firewood to gas and I changed. I have seen puppet show twice. It was good to see it as we used to see this kind of puppet dramas in my early childhood. And after years I am seeing like this…*..*ahhhh, hmmmm, I am becoming very old and I can remember only one thing from the puppet show that patients with chronic lung disease should visit the doctor regularly till their illness is cured*. (Female, in her 60’s)

*… I have learned a lot from the handouts and from the video that they (*health workers*) played on TV. Earlier, I didn’t know anything about the lung disease but now I am aware of it. I have also understood that how smoke can affect the lungs. Now I feel that I know something about it. Before, if someone has asthma, they feel ashamed to tell it out. Now, as you are coming as a group and putting up the puppet shows and video shows for the entire village, community started understanding that it’s a common disease and it will get healed soon. ….I have seen the change in the community. Those who have the disease and never wanted to say it out are coming out boldly and saying that they have this disease*. (Female, in her 40’s)

*They have put on the puppet show twice in our village, just opposite to my home but I didn’t understand. I think I didn’t care much so that I didn’t concentrate much*. (Female, in her 70’s)

..*After seeing the videos, my family is advising me to take inhaler before going to bed. They haven’t said these things before but after seeing the video only this change has happened. All these give happiness to me; …. Before they (*family*) never used to ask me, they have only seen me either crying or struggling to breathe and get air. Since they too watched the video shows along with me only this change has happened. (*Female, in her 40’s)

..It (handbills) *says about how we are getting the disease. If we breathe in smoke or if someone smokes, the smoke goes inside our lungs and it causes the disease. …. handout has pictures too for clear understanding*. (Female, in her 50’s)

…Hmmmm, *they (family) were not interested to watch the video messages. However, they have recognized now that I have a disease and they advise me not to do heavy work or eat cold fruits and vegetables*. (Female, in her 40’s)

.. *In our family, I haven’t observed any change in attitude after receiving the educational message. I have the disease and I have learned more about it. If they* (family members) *had the disease, they would have learned and changed*…*I haven’t noticed a change in the attitude of the community*. (male, in his 70’s)

## Discussion

This paper discusses the process of engaging diverse stakeholders including health care providers in the district, key people/gate keepers in the community and recipients of the intervention and their family members towards the development of a primary health care model for CRD in low resource settings in India and also their suggestions, opinions and feedback. The WHO recommends a people-centered, primary health care approach with a focus on community engagement as part of the Global action plan for effective prevention and management of non-communicable diseases including CRD.^15^

Studies report several barriers to primary health care based interventions at various levels of healthcare delivery: at the patient level, provider team level, organizational level, or policy level.^16^ A study undertaken in Sweden on stakeholders’ engagement in the prevention and management of Type 2 diabetes found community members were comfortable in living in close proximity with shared beliefs, values and resources. Authors identified several mismatches between awareness of patient needs and motivation to access available resources for type 2 diabetes (T2D) prevention. People were content to follow common, routine practices, however they were willing to make changes if their issues were addressed in a culturally appropriate manner ^17^ In addition, investigators observed the interaction between the communities and stakeholders was limited and concluded there are barriers in the collaboration between community, healthcare institutions and other stakeholders which will consequently affect the implementation of preventive interventions. Henceforth, the authors recommended exploring innovative ways to link the community to the healthcare sector and build the capacity of health systems for T2D prevention in socioeconomically disadvantaged communities. The identification of the influential stakeholders before starting the intervention is crucial and integral to its success. We recruited a panel of stakeholders that included patients and their family members, key community members and health care providers. Yoshita et al (2016) emphasized the significance of involving patients themselves, the most crucial stakeholders who are being impacted.^18^ However, the highlight of the literature has been the challenges associated with stakeholders’ participation in any new intervention.^19,20,21^ These challenges have not been empirically explored, especially within the context of low income countries.

This study is one of the few that discusses the definition of stakeholder engagement, processes involved in engaging them, their contribution to launching primary health care model of CRD management and various challenges in low-health literacy, socioeconomically disadvantaged settings in India. The qualitative research has shown that engaging the community and patients in risk alleviation, improving access to care pathway and preventive measures have great potential in enabling people to have increased control over the health choices they make. We found involving health workers, patients and their families in accessing the new intervention through education and training has paid rich dividends. Evidence from previous studies too suggests that community and clinical linkages established through health workers in the management of chronic diseases like HIV have significantly contributed to improved healthcare delivery and its cost-effectiveness.^22, 23^

The community engagement through educational interventions has shown mixed results in our study. It was difficult to develop appropriate tools acceptable to all. The investigators found that some of them will require a more personalised education. Muchiri et al (2019) in their study conducted in South Africa on stakeholders’ perceptions on diabetes management reported similar findings that the patients expressed desire for more personalised education, group meetings and sharing of testimonials by peers. They also suggested involvement of family members as one of the key strategies for positive behaviour change and concluded that adapting the interventions incorporating the participants’ choices for education methods need to be considered in future research.^24^

### Building Community trust

The initiative to involve the gate keepers (community advisory committee) in the community certainly helped to build a community trust. It was proved by the enthusiasm shown by them about the new CRD care initiative and their many suggestions including ways to inform the community, provision of community pharmacy managed by health workers and how to motivate people to comply with preventive measures. Studies have shown that community partnerships are most effective in building trust and thereby contributes significantly to health promotion and disease prevention programmes.^25,26, 27^

### Stakeholders’ engagement for sustainable delivery of evidence based care for CRD

For sustainable care, about 80 primary care physicians in the district were trained through CME, followed by a sample survey after 3 months to understand the outcome. The majority (70%) of the health care providers in the district, the backbone of health services, felt comfortable using mMRC and GINA guidelines for managing asthma and COPD respectively after attending the CME. About 42.9% of them changed the practice of using oral medications to recommended inhaler therapy. However, around 38% did not have access to spirometer or peak flow meters and a similar proportion of providers found it difficult to follow up the patients with clinical reviews at regular intervals. In low resource settings in India, most centers have no diagnostic facilities like spirometers and imaging facilities to make the correct diagnosis and recommend appropriate treatment. Culturally, most patients choose to access care from a randomly picked provider, making out-of pocket payment and often not returning to the same provider for follow-up that results in logistical difficulties in tracking disease progression and compliance to treatment in patients. In addition, on assessment of access to care, the patients opined that due to the chronic nature of the disease, medications should be delivered at the peripheral health centers (a community pharmacy) to avoid people losing a day’s wage by traveling 10-15 km to secondary care level hospitals and waiting hours for medication refills in the overcrowded busy outpatient department. Previous studies undertaken in other low resource settings around the world have identified similar barriers to accessing appropriate evidence based care for chronic diseases.^28,29,30^

### Impact of community engagement/ Community educational efforts

Knowledge deficits and beliefs concerning certain aspects of CRD and its treatment were revealed from both the patients’ and the community members’ accounts. These accounts also exposed disempowerment and uncertainty regarding access to inhalers, a mismatch between the awareness and access to evidence based care. It was enlightening for the investigators to see how the same education materials and methods were viewed differently by people of similar educational background and age. Some were able to understand the health messages from live puppet shows whereas some others did not engage. However the multiple educational methods and tools used for this research did raise the general awareness about CRD care as shown in the table.2. The knowledge deficits could hinder patients’ ability to make appropriate treatment choices and risk reduction efforts. The varying understanding of educational messages by the homogenous low literate population, delivered through popular methods and tools have not been widely explored in global health research context. It may offer a pragmatic solution to addressing adaptation of the new intervention.

**Table 2.** Feedback from the healthcare providers who attended the Continuing Medical Education workshop.

| Variable | Sub Variable | Frequency<br>(%)<br>n=21 |
| --- | --- | --- |
| **Profile of participants** |  |  |
| Gender | Female | 11 (52.4) |
|  | Male | 10 (47.6) |
| Current position | Assistant / Associate Professor | 2 (9.5) |
|  | Consultant | 5 (23.8) |
|  | General Practitioner | 14 (66.7) |
| Type of health care institution | Government | 8 (38.1) |
|  | Private | 12 (57.1) |
|  | Private & Government | 1 (4.8) |
| Average number of patients with chronic respiratory diseases seen per day | 1-5 | 9 (42.9) |
|  | 6-10 | 5 (23.8) |
|  | More than 10 | 6 (28.6) |
|  | Nil | 1 (4.8) |
| **Feedback** |  |  |
| Facing challenges to procure or access diagnostic tools like Peak flow meter, Spirometer | Slightly | 1 (4.8) |
|  | No | 12 (57.1) |
|  | Yes | 8 (38.1) |
| Able to conduct any health education on CRD | Irregular | 4 (19.0) |
|  | No | 6 (28.6) |
|  | Yes | 11 (52.4) |
| Facing challenges in the follow up review of patients | Occasionally | 6 (28.6) |
|  | No | 7 (33.3) |
|  | Yes | 8 (38.1) |
| Facing challenges to follow up CRD patients using mMRC guidelines for COPD and GINA guidelines for Asthma to optimize their management | Slightly | 1 (4.8) |
|  | No | 15 (71.4) |
|  | Yes | 5 (23.8) |
| Changes made in the daily practice | A more structured approach, with inclusion of PEFr and Spirometry | 2(9.5) |
|  | I have changed the use of oral medications to inhalers(MDI) | 9(42.9) |
|  | Patient education given on need for Spirometry, correct technique of inhaler use | 2(9.5) |
|  | Early diagnosis and intervention/Optimal management /quick assessment | 6(28.6) |
|  | Learned the indications for steroid inhalers and started using | 2(9.5) |

**Table 2.** **Comparing the change in attitude, subjective norms, perceived behavioural control, behavioural intention and behaviour related to CRD among participants (constructs of theory of planned behaviour based education) at the end of 1 year of intervention**

| Constructs of Theory of planned behaviour (Total score) | Mean Score (SD) |  |  |
| --- | --- | --- | --- |
|  | Baseline | End-line | Change(95% Confidence interval) |
| Attitude (45) | 30.30+3.16 | 35.33+4.51 | 5.0 (3.9, 6.2)* |
| Subjective norm (35) | 17.83+3.49 | 21.25+3.70 | 3.4 (2.4, 4.5)* |
| Perceived behavioural control(35) | 13.16±3.44 | 25.60±6.33 | 12.4 (10.9, 13.9)* |
| Behavioural Intention(40) | 23.29±3.02 | 23.10±4.60 | -0.08 (-1.0, 0.5) |
| Behaviour (5) | 2.51±.87 | 3.82±1.05 | 1.3 (1.0, 1.6)* |

\* Significant change

## Conclusion and implications

The findings of this study add to the literature on the numerous challenges faced by patients with CRD in low resource settings, indicating the need for identifying relevant stakeholders, continuous support of patients, personalised education, and capacity building of health care providers to navigate through the many hurdles faced by the patients. Overall, the community welcomed the idea of the new initiative to treat their diseases, however, the patients felt that they have limited capacity to procure inhalers and manage the cultural and economic factors that challenge CRD care. Given the significant challenges that the patients face, a feasible primary health care model needs to be developed incorporating strategies to deal with these challenges.

## Data Availability

All data produced in the present study are available upon reasonable request to the authors

## Acknowledgements

We are indebted to our research team at the study site who worked passionately to complete the study, frontline health care workers who volunteered to assist with the care, the RESPIRE team at the university of Edinburgh (lead partner) and other partner organisations for their support and suggestions and to all the participants who responded to stakeholders’ engagement activities.

## Supporting information

1. Fig 1. Stakeholders’ role to provide evidence based care for Chronic Respiratory Diseases
2. COREQ (COnsolidated criteria for REporting Qualitative research) Checklist

